# Accuracy Overstates Evidence Grounding and Abstention Reliability in Mammography Vision-Language Models

**DOI:** 10.64898/2026.09.10.26361944

**Authors:** Bowen Qu, Weixin Liu, Matthew Murrow, Matthew Burger, Xingyi Guo, Mihir Sachin Vaidya, Susannah L. Rose, Murat Kantarcioglu, Bradley A. Malin, Zhijun Yin

## Abstract

Answer accuracy alone cannot determine whether a Vision-Language Model (VLM) relies on clinically relevant mammographic evidence or recognizes when that evidence is unavailable. We introduce an evidence-grounded selective evaluation benchmark that evaluates Pathology classification and Abnormality identification together with label-aware lesion localization. Each original image–task instance is paired with a lesion-removal counterfactual and, when feasible, a size-matched non-lesion random-removal control. Lesion evidence is removed by local tissue reconstruction, while the random-removal view provides a task-irrelevant regional perturbation baseline of comparable spatial extent. We evaluate 16 general-purpose and medically specialized VLMs, including open-source and proprietary models, under a common protocol with the same prompt, image conditions, label spaces, and structured-output requirements. The results show substantial gaps between classification performance and evidence-grounded reliability. InternVL3.5-8B achieved the highest Pathology Macro-F1 at 53.75%, Huatuo Vision-34B achieved the highest Abnormality Exact Match at 52.44%, and GPT-5.6 Sol achieved the highest Grounded Answer Success at 21.99%. Lingshu-7B achieved the highest Counterfactual Specificity at 35.19%, whereas GPT-5.6 Sol achieved the highest Joint Reliability at 6.47%. These findings suggest that answer accuracy and abstention behavior can substantially overstate the reliability of current VLMs when predictions are not verified against the visual evidence on which they should depend.

## I. Introduction

Breast cancer is the most commonly diagnosed cancer and the leading cause of cancer-related death among women worldwide [1]. Mammography is a primary imaging modality for breast cancer screening and diagnostic assessment [2]. Accurate interpretation requires not only identifying findings such as masses, calcifications, architectural distortion, and asymmetry, but also assessing their characteristics and interpreting them across views, between breasts, and over time. These contextual comparisons are essential because a finding visible on one projection may be confirmed, recharacterized, or dismissed when the complete examination is considered.

Recent advances in medical Vision-Language Models (VLMs) have created opportunities to support mammography interpretation through interactive Visual Question Answering (VQA) [3], [4]. In this high-stakes setting, however, answer accuracy alone is insufficient. A reliable system should ground its prediction in the relevant image evidence, recognize when that evidence is insufficient, and abstain when the question cannot be supported by the available image.

Medical VQA benchmarks range from compact, expert-curated datasets such as VQA-RAD and SLAKE to large-scale collections such as PMC-VQA and OmniMedVQA [5]–[8]. These resources cover diverse imaging modalities, anatomical regions, and clinical questions, but they do not specifically focus on mammography. Mammography warrants separate studies because it generates low-contrast projection images in which overlapping breast tissue can obscure subtle masses, calcifications, and architectural distortions. MammoVQA provides a domain-specific benchmark by integrating 15 public mammography datasets for breast screening and diagnosis [9]. However, it focuses primarily on answer accuracy rather than whether a correct prediction is supported by the relevant lesion or changes when that lesion evidence is removed.

Related work examines this problem through medical hallucination evaluation, visual grounding, and selective prediction [10]–[14]. Medical hallucination evaluation tests whether a model produces claims that are unsupported by the image or clinical context; visual grounding tests whether an answer can be linked to the relevant image region; and selective prediction allows a model to abstain when the available evidence is insufficient. However, these capabilities are usually evaluated separately. This matters in mammography because diagnostic evidence often occupies a small, localized region. A correct label without matching lesion evidence may come from dataset priors rather than image interpretation. Conversely, a model that abstains may simply be conservative rather than sensitive to missing evidence. A joint evaluation of answer correctness, label-aware lesion grounding, and evidence-specific abstention under controlled evidence removal is therefore still needed.

We introduce an evidence-grounded selective VQA benchmark assembled from CBIS-DDSM, MIAS, and VinDr-Mammo [15]–[17]. The benchmark covers two complementary tasks. Pathology classification asks the model to predict the source-provided Benign or Malignant status associated with an annotated finding; this outcome label is distinct from a radiologist’s mammographic assessment. Multi-label Abnormality identification asks the model to identify all applicable findings among nine mammographic abnormality categories and recognize evidence for each prediction. For each original image–task instance, we create a lesion-removal counterfactual by replacing the annotated lesion region with reconstructed local breast tissue. When a valid control location exists, we also create a size-matched non-lesion random-removal control by editing a similarly sized lesion-free region while leaving the annotated lesion intact. The control helps determine whether a response change is specific to removal of lesion evidence or merely reflects sensitivity to image editing. All three views use the same question and task-specific label space. A reliable model should provide a correct, grounded answer on the original view, abstain after all lesion evidence required by the question has been removed, and preserve the correct answer on the random-removal control.

We evaluate four linked aspects of reliability across 16 general-purpose and medically specialized VLMs: whether each model answers correctly when lesion evidence is present; whether every predicted label is supported by a localized lesion; whether confidence and abstention distinguish reliable from unreliable responses; and whether the model abstains specifically after lesion evidence is removed while retaining the correct answer after a size-matched non-lesion edit. We report these behaviors separately and jointly for the same image–task instance. This design distinguishes correct but ungrounded answers, indiscriminate sensitivity to image editing, and evidence-specific model behavior.

## II. Related Work

### A. Medical and Mammography VQA Benchmarks

Medical VQA benchmarks span diverse imaging modalities and task formulations. VQA-RAD and SLAKE provide radiology-focused question–answer datasets [6], [18], PMCVQA scales medical VQA using biomedical figures and associated text [19], and OmniMedVQA integrates multiple sources to evaluate VLMs across modalities, anatomical regions, and question types [8]. These resources enable broad comparison of general-purpose and medical VLMs, but their heterogeneous tasks provide limited control over the specific visual evidence required for individual predictions.

Mammography-specific benchmarks address this gap in domain coverage. MammoVQA integrates 15 public datasets and more than 565,000 question–answer pairs covering breast density, abnormality identification, pathology classification, and BI-RADS assessment [9]. However, its evaluation primarily measures agreement with reference answers derived from source-dataset annotations and does not determine whether a correct prediction is supported by the corresponding lesion or remains valid when that evidence is removed.

### B. Evidence Grounding and Selective Reliability

Medical VLM reliability benchmarks increasingly evaluate behavior beyond answer accuracy. CARES examines factuality, uncertainty, fairness, safety, privacy, and robustness [20], while hallucination benchmarks use fake questions, mismatched image–question pairs, or altered answer sets to identify responses unsupported by the available input [10], [21]. Grounded medical VQA more directly links predictions to image evidence: HEAL-MedVQA provides doctor-annotated anatomical masks and evaluates region-aware reasoning under textual and visual perturbations [11], while A^3^Tune and CARE introduce region generation, attention alignment, or evidence– answer verification to strengthen visual grounding [22], [23]. These approaches nevertheless leave two aspects of evidence-based reliability insufficiently tested. First, localization of an anatomical region does not establish that the region supports the specific predicted lesion label. Second, a change in model output after regional perturbation does not establish evidence-specific reliance unless it is distinguished from general sensitivity to image editing. HEAL-MedVQA, for example, evaluates responses after replacing queried regions but does not explicitly test abstention or include a task-irrelevant regional control [11]. Our benchmark therefore uses label-aware grounding, requiring predicted evidence to match both the location and label of a supporting reference lesion.

### C. Counterfactual Evaluation and Abstention

Counterfactual interventions provide a complementary test of whether predictions depend on relevant visual evidence. Prior studies show that medical VLMs can retain apparently strong performance when informative image content is unavailable [24]; DrVD-Bench evaluates recognition of organs or lesions removed from edited images [25]; and recent work uses blank, shuffled, absent, or hard-negative images to quantify visual reliance and hallucination [26]. These interventions reveal shortcut behavior, but global substitutions alter both relevant and irrelevant visual information, while recognizing what was removed does not establish that a model appropriately revises its answer to the original task.

Selective prediction addresses the complementary question of whether a model withholds an answer when evidence is insufficient. Prior work studies selective VQA using risk– coverage analysis, response verification, and abstention-aware medical QA [13], [14], [27], but does not couple abstention with targeted removal of the visual evidence supporting the same prediction. Our benchmark combines these perspectives by pairing each original mammogram with a lesion-removal counterfactual and, when feasible, a size-matched non-lesion removal control. It therefore evaluates whether a model is correct, grounds its answer in the appropriate lesion, and selectively withholds that prediction when the supporting evidence is removed.

## III. Benchmark Design

Figure 1 provides an overview of the benchmark design, including the data sources, tasks, data curation process, and evaluation metrics, which are described in detail below.

**Fig. 1.**
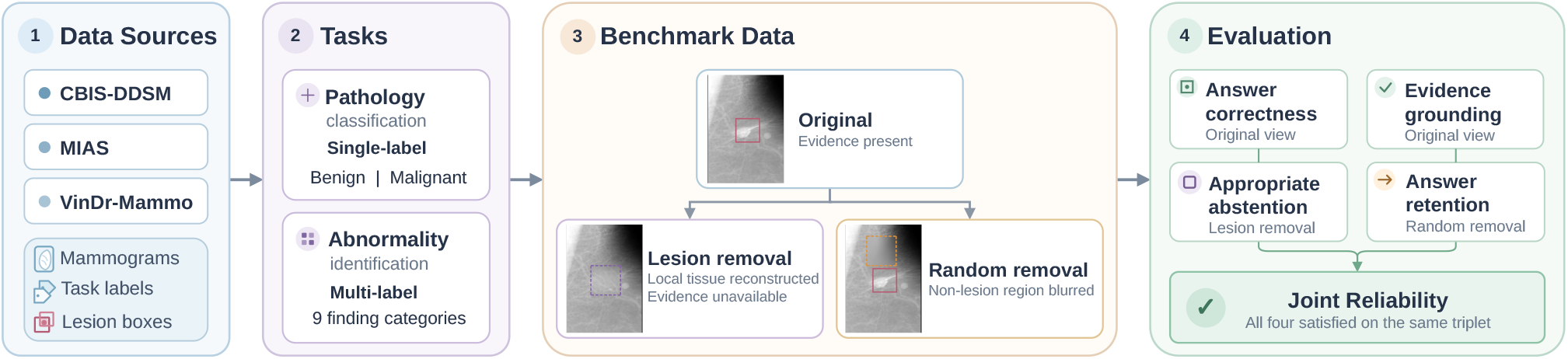
Overview of the evidence-grounded selective mammography VQA benchmark. Mammograms, task labels, and lesion boxes are assembled from CBIS-DDSM, MIAS, and VinDr-Mammo to support two tasks: single-label Pathology classification and multi-label Abnormality identification. Each original mammogram is paired with a lesion-removal counterfactual and, when feasible, a size-matched non-lesion random-removal control. Evaluation jointly measures answer correctness and label-aware evidence grounding on the original mammogram, appropriate abstention after lesion removal, and answer retention after random removal. Joint Reliability requires all four behaviors to be satisfied for the same task instance.

### A. Datasets and Task Definition

The benchmark selects MammoVQA records originating from CBIS-DDSM, MIAS, and VinDr-Mammo [9], [15]–[17]. CBIS-DDSM and MIAS contain digitized screen-film mammograms, whereas VinDr-Mammo contains full-field digital mammograms; none are synthesized two-dimensional images reconstructed from tomosynthesis. Each model input contains a single mammographic view rather than the complete multiview examination. Laterality and projection are not provided in the prompt. Some source images retain visible side or view markers, but their presence is inconsistent and no standardized side or view metadata are supplied.

We retain the unified MammoVQA labels rather than constructing a new clinical mapping. MammoVQA reports manually verifying its images, lesion annotations, and mapping of source labels to its unified answer space [9]; we preserve this mapping without additional clinical reinterpretation or independent radiologist review. The benchmark includes two tasks. Pathology classification predicts the source-provided *Benign* or *Malignant* status, derived from pathology-verified CBIS-DDSM labels or MIAS severity labels, rather than a new mammographic or BI-RADS assessment.

Abnormality identification requires all applicable labels from nine MammoVQA categories: *Calcification, Mass, Architectural distortion, Asymmetry, Miscellaneous, Nipple retraction, Suspicious lymph node, Skin thickening*, and *Skin retraction*. MIAS *CALC, CIRC/SPIC, ARCH, ASYM*, and *MISC* correspond to *Calcification, Mass, Architectural distortion, Asymmetry*, and *Miscellaneous*, respectively, while corresponding CBIS-DDSM and VinDr-Mammo labels retain their MammoVQA categories. The inherited Asymmetry label does not distinguish focal, global, or developing asymmetry, and MIAS *MISC* denotes other or ill-defined masses. Suspicious lymph node retains the VinDr-Mammo source annotation rather than being reassessed using newly defined criteria. Thus, these tasks measure agreement with inherited source labels from one view rather than complete clinical characterization. In both tasks, each predicted label must be linked to localized visual evidence.

### B. Image Conditions and Expected Responses

For each instance, the original mammogram is answerable because the annotated lesion evidence is present. In the lesion-removal condition, all annotated lesion regions required by the question are replaced with reconstructed local breast tissue; the edited image is therefore designated evidence-insufficient, and the expected response is abstention. It is not reassigned to *Normal*, because a local synthetic edit does not establish that the mammogram represents a genuinely normal examination. When feasible, the random-removal control blurs lesion-free breast-tissue regions matched one-to-one to the lesion-removal edit regions in width and height while leaving the annotated lesions intact; the expected behavior is therefore retention of the correct answer. The question, label space, answer mode, and prompt remain fixed across conditions, and the model is not informed of the image condition.

### C. Paired Counterfactual Construction

Paired counterfactual images test whether a prediction depends on the annotated lesion evidence. For an original image *I*_*i*_, let *ℬ*_*i*_ denote the set of annotated lesion boxes that constitutes the reference evidence, and Ω_*i*_ denote the corresponding edit regions derived from those boxes. The lesion-removal condition replaces the pixels in Ω_*i*_ with reconstructed local breast tissue. When feasible, the random-removal control blurs a set of lesion-free breast-tissue regions *ℛ*_*i*_ containing one control region for each lesion-removal edit region, with corresponding regions matched in width and height:

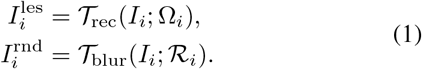

For lesion removal, we used a deterministic local tissue-reconstruction procedure motivated by exemplar-based inpainting and seamless image editing [28], [29]. Rather than attenuating the annotated lesion, the procedure reconstructs its local appearance from surrounding non-lesion breast tissue. Let *S*_*i*_ denote the surrounding non-lesion tissue used to estimate the local background. We first fit a robust quadratic surface,

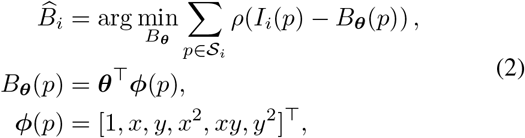

where *ρ* denotes the robust fitting objective. The fitted surface captures the low-frequency intensity variation of the local breast tissue. To restore fine-scale texture, we select a nonoverlapping non-lesion donor region with similar local intensity and texture statistics, following the general principle of patch-based correspondence used in exemplar-based image editing [28], [30]. When no suitable donor is available, the high-frequency residual is estimated from perilesional nonlesion tissue. We then define

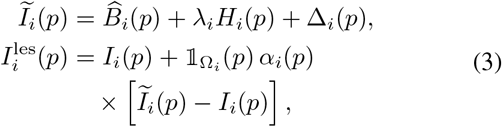

where 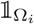 is the indicator of the edit region, *λ*_*i*_ matches the local texture scale, and Δ_*i*_ corrects low-frequency intensity differences along the edit boundary. The blending weight *α*_*i*_ provides a smooth transition between reconstructed and unmodified tissue, consistent with the general objective of seamless local image editing [29]. Specifically, *α*_*i*_(*p*) = *t*_*i*_(*p*)^2^(3*−*2*t*_*i*_(*p*)), where *t*_*i*_(*p*) is the normalized distance to the boundary of Ω_*i*_. Within the original, unpadded lesion region, *α*_*i*_(*p*) is set to one, ensuring that annotated lesion pixels are fully replaced rather than blended back into the image.

Each random control region lies within lesion-free breast tissue, avoids all annotated findings, and is matched one-toone to a lesion-removal edit region in width and height. Thus, the control matches region count and size, but not anatomical location or editing operator. This paired design tests whether a response changes specifically when annotated lesion evidence is removed, rather than as a generic reaction to editing a similarly sized non-lesion region.

### D. Evidence-Grounded Selective QA Protocol

All VLMs are tested using the same fixed prompt on all image conditions. Each model must rely only on visible mammographic evidence and return a structured response containing a complete answer set, an abstention decision, a normalized confidence estimate, label-aligned evidence, and a concise rationale. Each selected label must be paired with exactly one evidence region. When the visible evidence is ambiguous or insufficient, the model must abstain and return neither predicted labels nor evidence regions.

Evidence boxes use a shared normalized coordinate system. For an image of width *W* and height *H*, a pixel-space box (*x*_1_, *y*_1_, *x*_2_, *y*_2_) is mapped to a coordinate range with upper bound *S* as

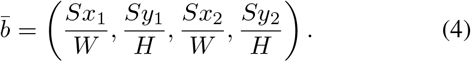

A box is valid only if it lies within the normalized range and has positive area. A strict parser checks response completeness, allowed labels, answer cardinality, consistency between the answer and abstention decision, confidence validity, and evidence geometry. Invalid responses are not repaired and receive no credit. The value of *S* is specified in Section IV-A, while the fixed prompt and structured response requirements are summarized in Table I.

**TABLE I.**
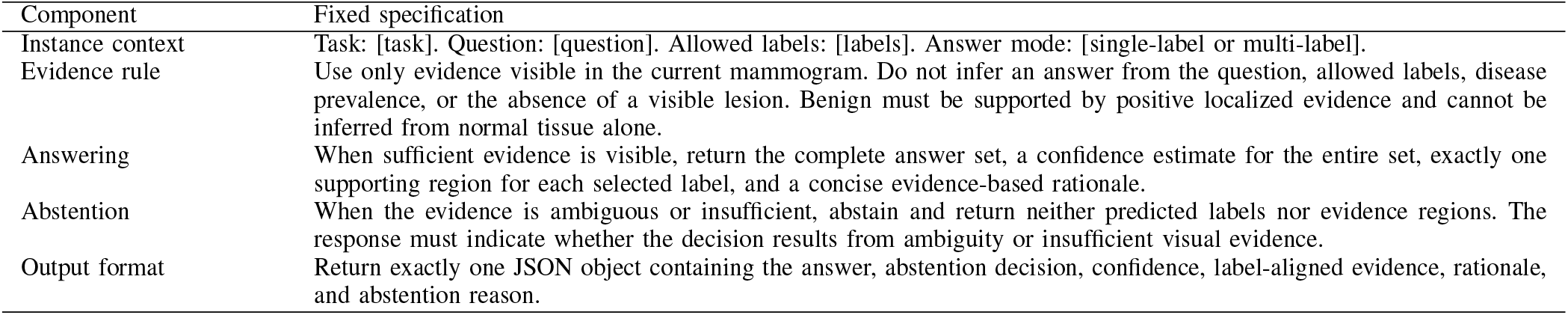
Fixed prompt and response protocol. Bracketed content is populated from each benchmark instance.

### E. Evaluation Metrics

The metrics are organized around four questions. Diagnostic metrics measure answer correctness on original mammograms. Grounded Answer Success tests whether a correct answer is accompanied by label-aligned lesion evidence. Coverage, Selective Accuracy, AURC, ECE, and Structured-output Validity characterize abstention, confidence behavior, and protocol compliance on original images. Counterfactual Specificity and Joint Reliability then evaluate whether model behavior changes specifically when lesion evidence is removed, with Joint Reliability requiring correctness, grounding, lesion-removal abstention, and random-control answer retention within the same triplet.

Response quality is evaluated on the original mammograms. Pathology classification is measured using macro-averaged F1 to reduce the effect of class imbalance. Abnormality identification is evaluated using *Exact Match*, which assigns credit only when the predicted label set is identical to the reference set with no missing or additional labels, together with macro-averaged F1 for performance across individual finding categories. Abstention on an original, answerable instance is counted as an incorrect prediction. Evidence grounding is evaluated using label-aware box matching. A predicted box for label *ℓ* may match only a reference region, and its grounding score is the maximum overlap with a compatible reference box. Let *C*_*i*_ indicate a correct complete answer on the original image, and let *V*_*i*_ indicate a valid one-to-one correspondence between the predicted labels and evidence regions. Intersection over Union (IoU) measures the overlap between a predicted evidence box and a compatible reference lesion box. Grounded Answer Success is defined as:

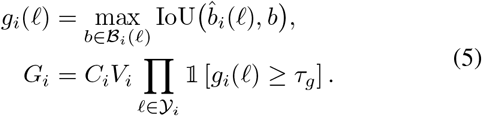

The benchmark reports the mean of *G*_*i*_ separately for Pathology and Abnormality. The grounding threshold *τ*_*g*_ and the evidence-validity criteria are specified in Section IV.

Selective prediction allows a model to abstain instead of returning a label when it considers the available evidence insufficient. On original mammograms, we summarize this behavior using *Coverage* and *Selective Accuracy. Coverage* is the proportion of all original instances receiving a strictly valid, non-abstaining response, while *Selective Accuracy* measures correctness among covered instances. The two metrics are interpreted jointly because excessive abstention can produce high Selective Accuracy.

Among strictly valid, non-abstaining responses, tie-aware area under the risk–coverage curve (AURC) summarizes how error risk changes as the confidence threshold is lowered. At each threshold, risk is the proportion of retained answers that are incorrect, and AURC integrates this risk across the attained coverage levels. Responses with identical confidence values enter the curve together, preventing an arbitrary ordering within ties; lower AURC indicates better confidence-based separation of correct and incorrect answers. We also report 10-bin equal-width Expected Calibration Error (ECE), the sample-frequency-weighted average of the absolute difference between mean confidence and empirical accuracy within each confidence bin; lower ECE indicates better calibration. Because AURC and ECE are computed only from covered responses, they are interpreted alongside overall Coverage. Structured-output Validity is the proportion of raw responses that pass all schema, allowed-label, answer-cardinality, confidence, abstention-consistency, and evidence-geometry checks.

It measures protocol compliance rather than clinical capability. Counterfactual evaluation includes only instances in *ℒ*_tri_, the set of instances with complete original, lesion-removal, and random-removal conditions. Let *N*_tri_ = |*ℒ*_tri_ |. For each *i ∈ ℒ*_tri_, let *A*_*i*_ indicate valid abstention after lesion removal and let *R*_*i*_ indicate preservation of the correct answer after random removal. *Counterfactual Specificity* (CS) measures evidence-sensitive behavior among instances answered correctly in the original condition, whereas *Joint Reliability* (JR) measures strict end-to-end success across all complete triplets:

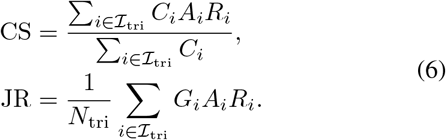

## IV. Experimental Setup

### A. Evaluation Data and Counterfactual Implementation

The final evaluation cohort contains 1,650 original image– task instances from 1,005 unique mammograms: 645 Pathology instances and 1,005 Abnormality instances. Of these instances, 1,250 are assembled from CBIS-DDSM, including 622 Pathology and 628 Abnormality instances; 46 are assembled from MIAS, with 23 instances per task; and 354 are assembled from VinDr-Mammo, all of which belong to the Abnormality task. The Pathology subset contains 377 Benign and 268 Malignant instances. The Abnormality subset contains 69 multi-label instances; label supports, which overlap for multilabel cases, are *Calcification* (375), *Mass* (580), *Architectural distortion* (22), *Asymmetry* (80), *Miscellaneous* (2), *Nipple retraction* (7), *Suspicious lymph node* (13), *Skin thickening* (18), and *Skin retraction* (2). The benchmark is used only for evaluation, and no benchmark records are used for model adaptation or model selection.

For each original instance, we construct the lesion-removal view in Eq. 1 using mask-aware local background reconstruction. The stored edit region is used when available; otherwise, the lesion box is expanded by 10% on each side and clipped to the image boundary. A robust quadratic surface fitted to surrounding non-lesion breast tissue estimates the local low-frequency background. High-frequency texture is drawn from a nearby, non-overlapping donor patch matched to the surrounding tissue statistics; when no suitable donor is available, a residual texture is synthesized from the perilesional support tissue. Boundary-intensity correction and soft feathering blend the reconstruction into the mammogram, while the original lesion box is fully replaced and pixels outside the edit region remain unchanged. Fixed reconstruction settings include a ring ratio of 0.40, a feather ratio of 0.08, 192 donor candidates, a search-radius ratio of 3.0, a minimum donor tissue fraction of 0.80, and a texture-blur ratio of 0.04.

This procedure yields 1,650 lesion-removal inputs and 1,577 matched random-removal controls. Each random region matches a lesion-removal region in size, remains within breast tissue, avoids annotated lesions and other control regions, and is blurred with a Gaussian radius of 10 pixels [31]. The breast-tissue threshold is the greater of 5 and the tenth percentile of positive grayscale intensities, and candidate regions require at least 70% tissue coverage. For 73 instances, no valid control satisfies all placement constraints; these instances remain in original- and lesion-removal analyses but are excluded from complete-triplet analyses. Automatic checks verify box geometry, image-boundary clipping, tissue coverage, non-overlap, and preservation of pixels outside edited regions. Box coordinates are normalized to [0, 1000] using Eq. 4.

We set the primary grounding threshold to *τ*_*g*_ = 0.30. Unlike detector-based localization, our benchmark evaluates evidence boxes generated zero-shot as part of a structured VLM response, where precise boundary optimization is not the objective. In addition, the reference annotations come from three datasets with differing annotation conventions and box granularity. We therefore use 0.30 as a permissive but meaningful criterion that requires spatial overlap with the relevant lesion while reducing sensitivity to annotation-boundary variation. This threshold is fixed across all models and tasks and is applied only after optimal one-to-one, label-aware matching, exact correspondence between answer and evidence labels, and successful matching of every predicted evidence item. Predicted boxes are rejected if their normalized area ratio is at most 0.0005 or at least 0.25, or if they are edge-degenerate; edge degeneracy denotes a box that touches the image boundary within one normalized coordinate unit and is additionally tiny or has an aspect ratio of at least 10. Thresholds of 0.40 and 0.50 are reported as stricter sensitivity analyses that place progressively greater emphasis on boundary agreement.

### B. Evaluated Models

We evaluate 16 VLMs, including general-purpose and medically specialized models with open-weight or commercial access. The general-purpose open-weight models are Qwen2.5-VL-7B, Qwen2.5-VL-72B, Qwen3-VL-32B, LLaVA-NeXT-7B, InternVL3.5-8B, and InternVL3-78B. The medically specialized open-weight models are LLaVA-Med-7B, Huatuo Vision-7B, Huatuo Vision-34B, Huatuo-Qwen-7B, MedGemma-4B, MedGemma-27B, Lingshu-7B, and Aloe Vision-72B. The commercial models are GPT-5.6 Sol and Gemini 3.5 Flash. Among the models for which parameter counts are disclosed, sizes range from 4B to 78B. All models receive identical test inputs and response requirements.

### C. Inference and Output Processing

Table I summarizes the prompt specification. Model-specific adapters handle only the image formatting and message serialization required by each interface. Open-weight models are served through a common local inference framework, whereas commercial models are accessed through provider APIs. We use a temperature of 0, a nucleus sampling parameter of 1.0, and a maximum output length of 512 tokens, with hidden-reasoning modes disabled when possible or set to the minimum supported level. One response is generated per input. Provider-side structured-output controls are used only to constrain response syntax. All outputs are subsequently evaluated by the same benchmark-side parser, which verifies JSON structure, allowed labels, answer cardinality, confidence values, abstention consistency, and evidence validity. Invalid outputs receive no credit and are not repaired.

### D. Statistical Analysis

We report 95% confidence intervals obtained from 1,000 grouped nonparametric bootstrap resamples with a random seed of 42, keeping all tasks and views from the same patient/study group together. For task-specific metrics, we resample the patient/study groups represented in the corresponding task. For counterfactual metrics, we resample only groups with complete original, lesion-removal, and random-removal triplets. The interval bounds correspond to the 2.5th and 97.5th percentiles and are reported for primary metrics and key paired comparisons. For each paired comparison, the metric difference is computed within every resample. A difference is considered statistically supported only when the resulting interval excludes zero.

## V. Results

### A. Performance on Original Mammograms

Table II reports model performance on the 1,650 original image–task instances. We retain all original instances in the denominator and count abstentions and invalid outputs as incorrect. Figure 3 and the final subsection consider Coverage and Selective Accuracy together. The table also reports tie-aware AURC and 10-bin equal-width ECE; both require interpretation alongside Coverage.

**TABLE II.** Diagnostic performance and confidence diagnostics on original mammograms. **P F1** denotes Pathology Macro-F1, **A EM** denotes Abnormality Exact Match, **A F1** denotes Abnormality Macro-F1, and **v****alid** denotes strict structured-output validity. **Aurc** denotes the tie-aware area under the risk–coverage curve, and **Ece** denotes 10-bin equal-width expected calibration error. Both are computed from strictly valid, non-abstaining responses to original mammograms. All values are percentages. Underlining marks the best result within each model block, and boldface also marks the overall best result for the three diagnostic metrics. Valid, AURC, and ECE are diagnostic measures rather than standalone ranking metrics. Lower AURC and ECE values are meaningful only at comparable levels of Coverage.

| Model | P F1 | A EM | A F1 | Valid | AURC | ECE |
| --- | --- | --- | --- | --- | --- | --- |
| Qwen2.5-VL-7B | 36.44 | 27.16 | 6.14 | 86.67 | 52.94 | 28.21 |
| LLaVA-NeXT-7B | 36.89 | 32.54 | 6.04 | 100.00 | 57.33 | 42.67 |
| InternVL3.5-8B | <u>53.75</u> | 20.80 | 9.26 | 98.61 | 59.57 | 54.86 |
| MedGemma-4B | 33.71 | 20.20 | 9.79 | 92.67 | 67.18 | 60.91 |
| LLaVA-Med-7B | 0.00 | 2.49 | 1.64 | 6.91 | 78.07 | 21.93 |
| Huatuo Vision-7B | 36.89 | <u>52.34</u> | 12.99 | 99.88 | 43.19 | 41.26 |
| Huatuo-Qwen-7B | 38.37 | 22.79 | <u>13.58</u> | 92.79 | 58.14 | 45.11 |
| Lingshu-7B | 17.84 | 24.98 | 8.09 | 100.00 | 54.26 | 47.94 |
| Qwen3-VL-32B | 16.10 | 35.02 | 13.07 | 100.00 | 55.40 | 43.65 |
| Qwen2.5-VL-72B | 4.18 | 18.81 | 6.87 | 90.18 | 44.49 | 26.37 |
| InternVL3-78B | 31.44 | 31.04 | 11.56 | 90.85 | 55.87 | 47.22 |
| MedGemma-27B | <u>47.66</u> | 28.16 | 9.85 | 100.00 | 60.78 | 43.25 |
| Huatuo Vision-34B | 45.90 | **52.44** | <u>15.20</u> | 99.94 | 45.12 | 33.46 |
| Aloe Vision-72B | 36.89 | 0.80 | 7.63 | 48.97 | 63.59 | 42.80 |
| GPT-5.6 Sol | <u>52.72</u> | <u>47.26</u> | **23.19** | 100.00 | 37.76 | 31.23 |
| Gemini 3.5 Flash | 52.05 | 41.49 | 16.33 | 99.94 | 40.25 | 36.57 |

**TABLE III.** Grounded Answer Success on original mammograms. A successful instance requires a complete correct answer and valid label-aware evidence that satisfies the primary grounding criterion for every answer label. All values are percentages. Underlining marks the best result within each model block while boldface marks the best result overall.

| Model | Grounded Answer Success |  |
| --- | --- | --- |
|  | Pathology | Abnormality |
| Qwen2.5-VL-7B | 0.31 | 0.60 |
| LLaVA-NeXT-7B | 0.00 | 0.00 |
| InternVL3.5-8B | <u>4.81</u> | <u>3.18</u> |
| MedGemma-4B | 0.62 | 0.40 |
| LLaVA-Med-7B | 0.00 | 0.00 |
| Huatuo Vision-7B | 1.09 | 0.70 |
| Huatuo-Qwen-7B | 1.09 | 0.80 |
| Lingshu-7B | 1.09 | 2.39 |
| Qwen3-VL-32B | 4.96 | <u>12.94</u> |
| Qwen2.5-VL-72B | 0.16 | 0.60 |
| InternVL3-78B | 1.40 | 0.90 |
| MedGemma-27B | 0.62 | 0.60 |
| Huatuo Vision-34B | 0.00 | 0.00 |
| Aloe Vision-72B | 0.78 | 0.00 |
| GPT-5.6 Sol | **21.86** | **21.99** |
| Gemini 3.5 Flash | 18.29 | 21.59 |

**TABLE IV.** Counterfactual reliability across complete original, lesion-removal, and random-removal triplets. Counterfactual Specificity is the proportion of correct original answers for which a model validly abstains after lesion removal and retains the correct answer after random removal. Joint Reliability additionally requires valid label-aware grounding on the original image and uses all complete triplets as the denominator. All values are percentages. Underlining marks the best result within each model block, and boldface also marks the best result overall.

| Model | Counterfactual Specificity | Joint Reliability |
| --- | --- | --- |
| Qwen2.5-VL-7B | 1.21 | 0.00 |
| LLaVA-NeXT-7B | 0.00 | 0.00 |
| InternVL3.5-8B | 0.00 | 0.00 |
| MedGemma-4B | 6.83 | 0.13 |
| LLaVA-Med-7B | 0.00 | 0.00 |
| Huatuo Vision-7B | 0.00 | 0.00 |
| Huatuo-Qwen-7B | 0.00 | 0.00 |
| Lingshu-7B | <u>35.19</u> | <u>1.08</u> |
| Qwen3-VL-32B | 16.54 | 2.98 |
| Qwen2.5-VL-72B | <u>30.69</u> | 0.25 |
| InternVL3-78B | 11.46 | 0.06 |
| MedGemma-27B | 0.49 | 0.00 |
| Huatuo Vision-34B | 4.47 | 0.00 |
| Aloe Vision-72B | 0.00 | 0.00 |
| GPT-5.6 Sol | 22.70 | **6.47** |
| Gemini 3.5 Flash | <u>23.42</u> | 5.07 |

No model ranked first on all three diagnostic metrics. InternVL3.5-8B had the highest Pathology Macro-F1 at 53.75%, followed by GPT-5.6 Sol at 52.72% and Gemini 3.5 Flash at 52.05%. The 95% confidence interval for the paired difference between InternVL3.5-8B and GPT-5.6 Sol was [*−*4.03, 6.44] percentage points, and did not support a reliable difference between the two models.

For Abnormality identification, Huatuo Vision-34B and Huatuo Vision-7B had the two highest Exact Match scores, at 52.44% and 52.34%, respectively. The 95% confidence interval for their paired difference was [*−*2.39, 2.69] percentage points. GPT-5.6 Sol, however, had the highest Abnormality Macro-F1 at 23.19%, exceeding Gemini 3.5 Flash by 6.86 percentage points, with a paired interval of [2.37, 11.65]. The different Exact Match and Macro-F1 rankings show that high complete-set accuracy does not necessarily reflect balanced performance across individual abnormality categories. Among the three categories with at least 80 positive instances, the highest category-level F1 values achieved by any model were 72.54% for Mass (Huatuo Vision-34B), 63.76% for Calcification (GPT-5.6 Sol), and 17.01% for Asymmetry (Huatuo-Qwen-7B); the other six categories each contained at most 22 positives and were too sparse for stable model comparisons. Structured-output validity ranged from 6.91% for LLaVA-Med-7B to 100.00% for several models. All models other than LLaVAMed-7B and Aloe Vision-72B exceeded 86% validity. This measure reflects adherence to the response protocol and is not treated as independent evidence of clinical capability.

### B. Evidence Grounding on Original Mammograms

We next ask whether correct answers are supported by the appropriate lesion evidence. GPT-5.6 Sol had the highest Grounded Answer Success on both tasks, with 21.86% for Pathology and 21.99% for Abnormality. Gemini 3.5 Flash ranked second at 18.29% and 21.59%, respectively. The paired difference for Pathology was 3.57 percentage points, with a 95% confidence interval of [0.31, 6.82]. For Abnormality, the difference was 0.40 percentage points, with an interval of [*−*2.59, 3.18].

Qwen3-VL-32B was the strongest open-source model for grounding, reaching 4.96% for Pathology and 12.94% for Abnormality. GPT-5.6 Sol still exceeded Qwen3-VL-32B on Abnormality by 9.05 percentage points, with a paired interval of [6.37, 11.54]. No model exceeded 22% on either task, so correct answers were rarely accompanied by sufficiently aligned lesion evidence.

Grounding changes the ranking from the diagnostic results in Table II. Huatuo Vision-34B reached 52.44% Abnormality Exact Match, but none of its responses satisfied the primary grounding criterion. Huatuo Vision-7B showed a similar pattern, with 52.34% Exact Match and only 0.70% Grounded Answer Success. InternVL3.5-8B had the highest Pathology Macro-F1 but only 4.81% Grounded Answer Success. Answer correctness alone therefore does not show that a prediction is supported by the relevant lesion evidence.

The threshold-sensitivity analysis supports this conclusion. Across both tasks, the highest Grounded Answer Success decreased from 21.99% at *τ*_*g*_ = 0.30 to 17.91% at *τ*_*g*_ = 0.40 and 15.42% at *τ*_*g*_ = 0.50. GPT-5.6 Sol and Gemini 3.5 Flash exchanged order for some task–threshold combinations, indicating that their fine-grained ranking depends on the required degree of boundary agreement. Qwen3-VL-32B nevertheless remained the strongest open-source model on both tasks at all three thresholds. Thus, stricter thresholds reduce the absolute success rates and can affect closely ranked models, but they do not alter the central finding that correct answers are seldom accompanied by consistently aligned lesion evidence.

Figure 2 shows the difference between answer correctness and evidence localization in two selected cases. GPT-5.6 Sol and Qwen3-VL-32B answer both cases correctly and satisfy the localization criterion. Huatuo-Qwen-7B also returns the correct labels but places the predicted evidence away from the reference lesions. The multi-label case also shows that grounding must be checked separately for every answer label.

**Fig. 2.**
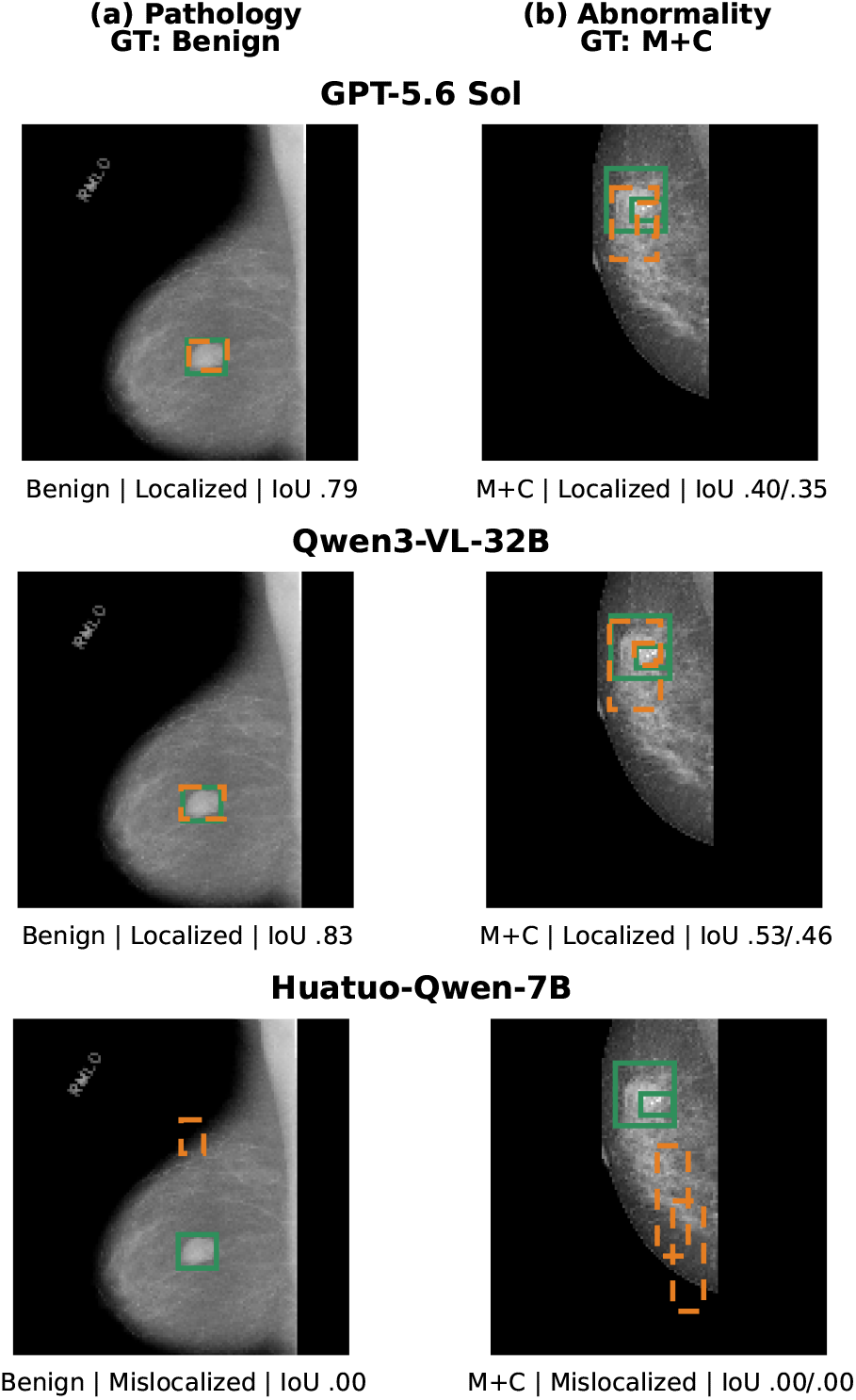
Examples of label-aware grounding for Pathology (left) and multilabel Abnormality (right). Green solid boxes indicate reference evidence, and orange dashed boxes indicate model-predicted evidence. “Localized” requires IoU *≥ τ*_*g*_ = 0.3; “Mislocalized” denotes a correct answer for which the predicted evidence fails this criterion. M and C denote Mass and Calcification, respectively, and paired IoUs are reported in M/C order. These cases show different grounding behaviors and do not represent aggregate model rankings.

**Fig. 3.**
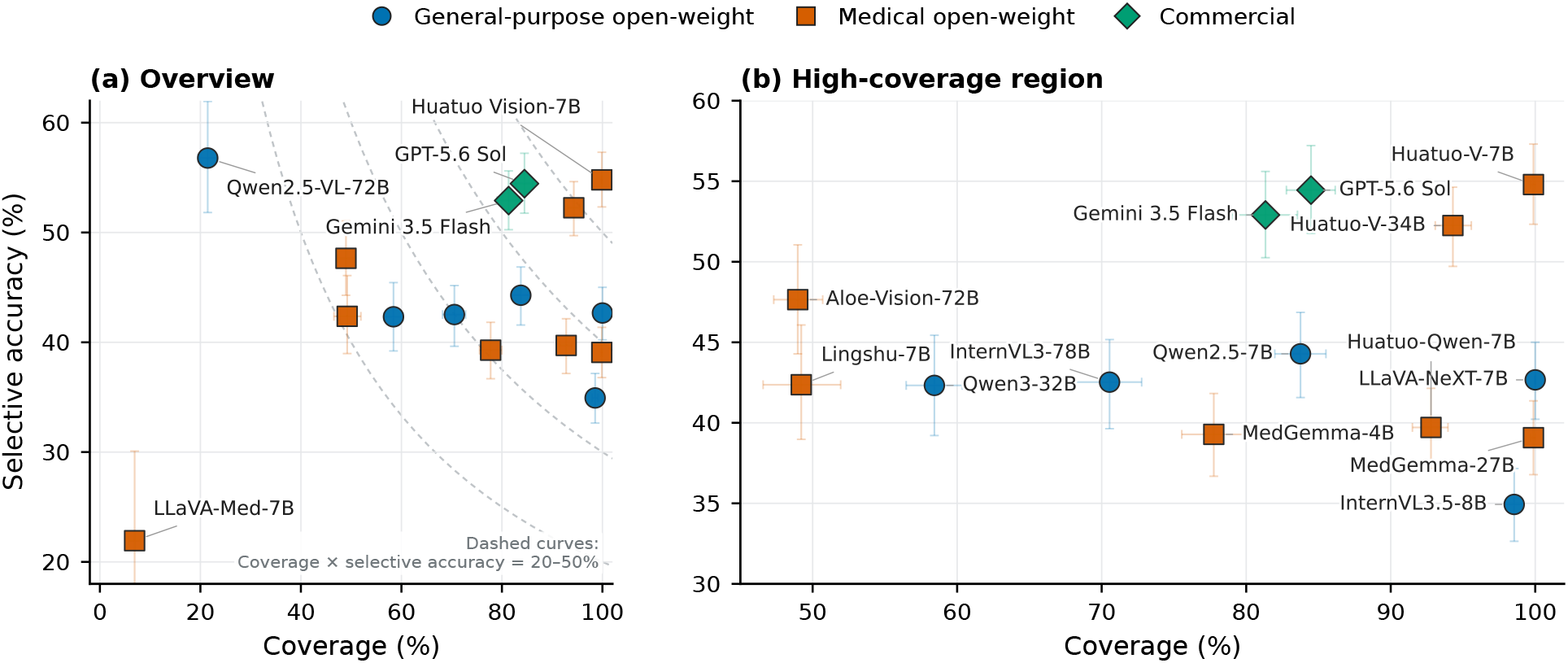
Selective operating points on original mammograms. Panel (a) shows the full operating space, and panel (b) enlarges the dense high-coverage region. Each point represents one model. Color and marker shape distinguish general-purpose open-source, medical open-source, and proprietary models. Coverage is the proportion of all original instances receiving a strictly valid, non-abstaining response, whereas Selective Accuracy is correctness among covered instances. Horizontal and vertical error bars show marginal 95% confidence intervals for Coverage and Selective Accuracy, respectively, estimated from 1,000 grouped bootstrap resamples, keeping all tasks and views from the same patient/study group together.

### C. Selective Prediction and Counterfactual Reliability

Before turning to lesion removal, we examine how often each model returns a strictly valid, non-abstaining response when the required evidence is present. Figure 3 plots the resulting operating points on original mammograms. Coverage is the proportion of all original instances receiving a strictly valid, non-abstaining response, whereas Selective Accuracy is correctness among covered instances.

For visual reference, panel (a) also shows contours of constant covered-and-correct rate, representing the proportion of all original instances for which a model produces a valid, non-abstaining, and correct response. For example, models on the 40% contour achieve this outcome on 40% of all instances, despite potentially differing in Coverage and Selective Accuracy. These contours are intended only as descriptive guides and are not used as an additional ranking metric.

Figure 3 shows several patterns. Qwen2.5-VL-72B had the highest point estimate of Selective Accuracy at 56.78% but covered only 21.45% of the original instances. Its Coverage and Selective Accuracy correspond to a covered-and-correct rate of 12.18%, below the 20% reference contour. The structured-output validity of 90.18% in Table II suggests that the low Coverage mainly reflects abstention rather than outputformat failure. LLaVA-Med-7B differs in that its low Coverage coincides with structured-output validity of only 6.91%.

Huatuo Vision-7B is closest to the upper-right corner, with 99.88% Coverage and 54.79% Selective Accuracy, corresponding to a covered-and-correct rate of 54.72% and placing it above the 50% reference contour. GPT-5.6 Sol and Gemini 3.5 Flash have intermediate operating points. Their Coverage values are 84.48% and 81.33%, and their Selective Accuracy values are 54.45% and 52.91%, respectively. These operating points correspond to covered-and-correct rates of 46.00% for GPT-5.6 Sol and 43.03% for Gemini 3.5 Flash, placing both between the 40% and 50% reference contours. The vertical confidence intervals overlap substantially, so the small differences in Selective Accuracy do not establish a clear ranking.

The confidence diagnostics in Table II give a similar picture. We computed tie-aware AURC and 10-bin equal-width ECE among strictly valid, non-abstaining responses to original mammograms. Lower values are better for both measures, but the values must be interpreted alongside Coverage. GPT-5.6 Sol had the lowest AURC at 37.76% (95% CI: 34.33–41.22%), followed by Gemini 3.5 Flash at 40.25% (36.60–43.63%). Their ECE values remained high at 31.23% (28.47–33.71%) and 36.57% (33.98–39.18%), respectively. The lowest ECE, 21.93%, came from LLaVA-Med-7B, but this value is difficult to interpret because the model covered only 6.91% of the original instances and used a single confidence level. Qwen2.5-VL-72B likewise had an ECE of 26.37% but used only three confidence levels and covered 21.45% of the original instances. Several other models also produced constant or highly discrete confidence values. Self-reported confidence therefore provided limited calibration information and did not resolve the Coverage–accuracy trade-off.

These original-view operating points alone cannot show whether the relevant lesion drives model answers or abstentions. Qwen2.5-VL-72B combines high Selective Accuracy with broad abstention, while Huatuo Vision-7B combines a strong upper-right operating point with very low Grounded Answer Success. The lower Coverage of Gemini 3.5 Flash relative to GPT-5.6 Sol also does not translate into higher Joint Reliability. We test this relationship directly using the paired lesion-removal and random-removal views.

Counterfactual Specificity produced a different model ordering from the original-view analyses. Lingshu-7B achieved the highest point estimate at 35.19%, followed by Qwen2.5-VL-72B at 30.69%. Their 4.50 percentage point difference was not statistically resolved, with a paired 95% confidence interval of [*−*2.61, 11.43]. Gemini 3.5 Flash and GPT-5.6 Sol followed at 23.42% and 22.70%, respectively. The two highest point estimates were therefore obtained by open-weight models, showing that strong lesion-removal sensitivity was not confined to commercial systems.

Joint Reliability produced a different ordering. GPT-5.6 Sol had the highest value at 6.47%, followed by Gemini 3.5 Flash at 5.07%. Their 1.40 percentage point difference had a paired 95% confidence interval of [*−*0.06, 3.03], which included zero. Qwen3-VL-32B had the highest Joint Reliability among the open-weight models at 2.98%. GPT-5.6 Sol exceeded Qwen3-VL-32B by 3.49 percentage points, with a paired interval of [2.00, 5.06]. Even the strongest models therefore satisfied the complete criterion on only a small fraction of triplets.

The component measures illustrate why high counterfactual sensitivity did not translate directly into end-to-end reliability. Lingshu-7B validly abstained on 64.12% of lesion-removal inputs and achieved 35.19% Counterfactual Specificity, yet its Joint Reliability was only 1.08%. Qwen2.5-VL-72B abstained on 73.21% of lesion-removal inputs and reached 30.69% Counterfactual Specificity, but low original-view Coverage and weak grounding limited Joint Reliability to 0.25%. By contrast, Huatuo Vision-7B answered nearly every original instance but abstained after lesion removal in only 0.61% of cases, yielding 0.00% Counterfactual Specificity and no jointly reliable triplets. Across all models, diagnostic answering, evidence grounding, counterfactual sensitivity, and Joint Reliability remained only weakly aligned.

## VI. Discussion and Conclusion

Our findings are broadly consistent with recent medical VQA benchmarks, but they also reveal a gap between several forms of reliability that are often evaluated separately. OmniMedVQA found that medical specialization does not consistently improve medical VQA performance, while MammoVQA showed that both general-domain and medical VLMs remain unreliable for mammography interpretation [8], [9]. We observe a similar pattern: neither model scale nor medical specialization yields consistent gains across diagnostic accuracy, grounding, and selective behavior. The discrepancy between answer correctness and Grounded Answer Success also supports the finding of HEAL-MedVQA that medical VLMs can produce correct answers while relying on weak or irrelevant visual evidence [11]. Our counterfactual evaluation adds a further distinction. Models with high Counterfactual Specificity do not necessarily achieve high Joint Reliability, and the two highest Counterfactual Specificity estimates come from open-weight models, whereas proprietary models achieve the strongest original-view grounding and Joint Reliability. This result is consistent with the broader observations of CARES and MedHEval that task-level performance alone is insufficient to establish trustworthy or robust medical VLM behavior [20], [21]. These results indicate that answer correctness, lesion localization, sensitivity to evidence removal, and calibrated abstention capture different aspects of model reliability and should be evaluated jointly.

Several limitations motivate future work. First, our evaluation is limited to single-view mammograms; extending the benchmark to complete examinations could incorporate bilateral and multiview comparison, prior studies, and complementary imaging such as tomosynthesis or ultrasound. This would test whether grounding and abstention improve with the broader context used in clinical interpretation. Second, larger and more balanced datasets with breast density labels would enable subgroup analyses across lesion types and clinically relevant patient characteristics. Third, future work could directly assess rationale faithfulness and whether generated explanations align with localized evidence. Finally, prospective studies are needed to determine whether evidence-grounded and abstention-aware behavior improves radiologist-assisted interpretation and other realistic clinical workflows.

## Data Availability

The data analyzed in this study were obtained through MammoVQA and originate from CBIS-DDSM, MIAS, and VinDr-Mammo. All data were publicly available before the initiation of the study. Access information and citations for the datasets are provided in the manuscript and through the links below. No newly collected or identifiable patient data were used.

https://github.com/PiggyJerry/MammoVQA

https://www.cancerimagingarchive.net/collection/cbis-ddsm/

https://www.repository.cam.ac.uk/items/b6a97f0c-3b9b-40ad-8f18-3d121eef1459

https://physionet.org/content/vindr-mammo/1.0.0/

## VII. Funding Acknowledgment

This research was, in part, funded by the Advanced Research Projects Agency for Health (ARPA-H). The views and conclusions contained in this document are those of the authors and should not be interpreted as representing the official policies, either expressed or implied, of the United States Government.

